# Determining the optimal volume of reactive balance training after stroke – a pilot randomized controlled trial

**DOI:** 10.64898/2026.09.02.26362056

**Authors:** Avril Mansfield, David Jagroop, Cynthia J Danells, Tanvi Bhatt, Elizabeth L. Inness

## Abstract

**Background:** Our long-term goal is to determine the optimal volume of reactive balance training (RBT) in people with sub-acute stroke. This study aims to inform the design of a larger trial to address this long-term goal.

**Trial design:** Assessor-blinded three-arm pilot randomized controlled trial.

**Methods:** Participants (n=36) with sub-acute stroke were randomly allocated to one of three groups: 1, 3, or 6 RBT sessions. All sessions were delivered over 2-3 weeks. Feasibility outcomes included recruitment and retention rates, intervention fidelity and adherence, and completeness of outcome assessments. Participants reported falls and physical activity for 6 months post-training; falls data were used to estimate sample size for a future trial. Functional balance, balance confidence, and balance reactions were assessed at: 1) study enrolment; 2) post-training; and 3) 6 months post-training to estimate effect sizes for a future trial.

**Results:** Thirty-five participants initiated training and 34 contributed post-training assessment data. Intervention adherence was high: participants completed 1.0, 2.9, and 5.7 RBT sessions on average in the 1-, 3- and 6-session, respectively. Functional and questionnaire outcomes were ≥80% complete at most time points, although follow-up questionnaires at 2 and 4 months met only 74% completeness. Falls monitoring data were available for 30 participants, with 15 falls reported. Estimated sample sizes for a definitive trial using falls as the primary outcome (n=408-782) were not feasible for a single site; alternative primary outcomes (stability following balance perturbations) produced more feasible sample size estimates (n=30-72). Minor adverse events were reported by 1, 2, and 6 participants in the 1-, 3- and 6-session groups, respectively.

**Conclusions:** The modified intervention and assessment procedures were feasible; however, a definitive single-site trial using falls as the primary outcome is not feasible. Stability-based outcomes may support a more feasible future trial.

**Trial registration:** clinicaltrials.gov, NCT04219696

## INTRODUCTION

Reduced balance and mobility are common after stroke, contributing to increased risk of falling in daily life.^1,2^ Reactive balance training (RBT) is an emerging balance training approach that aims to improve reactive balance control; clients experience repeated balance perturbations (or loss of balance) to practice and improve control of balance reactions, like reactive stepping.^3^ There is good evidence that RBT can improve reactive balance control^4,5^ and prevent falls in daily life.^3^

There is wide variability in the training approaches used in previous RBT studies. For example, one systematic review found that the total duration of RBT programs in clinical trials ranged from 15 minutes in a single session to 24 hours spread over several weeks.^6^ It is possible that reactive balance control can improve with a low volume of RBT. For example, among healthy older adults, just 24 balance perturbations within a single RBT session led to lasting improvements (i.e., 6-12 months) in reactive balance control,^7^ and reduced rate of falls in daily life.^8^ Among people with chronic stroke, a single session of RBT was associated with improved reactive balance control at 3 weeks^9^ and 6 months^10^ post-training. In contrast, secondary analysis of data from a randomized controlled trial suggested that greater improvements in reactive balance control were observed for those who completed a higher volume of training.^11^ Therefore, the optimal volume of RBT post-stroke still remains unclear.

Physiotherapists also report that they would like to know the optimal training parameters for RBT; e.g., the “right amount” of training.^12^ Healthcare professionals often struggle to implement new practices due to competing priorities, and physiotherapists note that limited time with their clients is a barrier to implementing RBT.^13,14^ Therefore, knowing the minimum number of sessions required to show improvements in reactive balance control may facilitate implementing it in practice.

The long-term goal of this work is ***to determine the optimal volume of RBT in people with sub-acute stroke***. This assessor-blinded pilot randomized controlled trial (RCT) aimed ***to inform the design of a larger trial to address this long-term goal***. Specifically, we aimed to answer the following research questions about the larger trial with this pilot study: 1) what is the optimal sample size, using falls in daily life as a primary outcome; 2) how long will it take to achieve this sample size; 3) are the proposed secondary outcome measures feasible (i.e., ≥80% complete at each time point); 4) how feasible is it to prescribe a specific volume of RBT to people with sub-acute stroke within routine out-patient rehabilitation (i.e., mean number of sessions attended is ≥75%); and 5) what two intervention groups should be included in the larger trial?

## METHODS

### Patient and public involvement

This study was designed without patient involvement. Patients were not invited to comment on the study design and were not consulted to develop patient relevant outcomes. Some trial design elements were informed by participant feedback from a previous study.^4^ Patients were not invited to contribute to the writing or editing of this document for readability or accuracy.

### Trial design and changes to trial protocol

This study was a parallel-group assessor-blinded three-arm pilot superiority RCT, with 1:1:1 group allocation. Research ethics approval was received from the Research Ethics Board of the University Health Network (Study ID: 19-6001, initial approval 17 January 2020). The trial is reported according to the primary CONSORT statement,^15^ and the CONSORT extension for pilot and feasibility studies.^16^ The way that the intervention was delivered was modified in response to low recruitment, as detailed below. The research team collectively decided on any protocol amendments, which were approved by the institutional Research Ethics Board prior to implementation.

### Trial setting

This study took place at the Toronto Rehabilitation Institute, University Health Network. This facility provides specialized in-and out-patient stroke rehabilitation to individuals in the sub-acute stage of stroke recovery

### Eligibility criteria

Participants were people with sub-acute stroke (<6-months post-stroke). Participants were eligible if they: 1) could stand independently for >30s; 2) could walk without assistance of another person for >10m; and 3) were living in the community. Participants were excluded if they had:

- Completed RBT during in-patient rehabilitation;
- Lower extremity amputation, weight-bearing restrictions, recent lower-extremity injury or surgery (e.g., fracture), acute back or lower-limb pain, halo, aspen collar, history of fragility fracture and/or severe osteoporosis/osteopenia, contractures that prevent neutral hip or ankle;
- Activity restrictions following cardiac event/surgery, abnormal or unstable cardiovascular responses to exercise, arterial dissection;
- Severe spasticity in the legs;
- Cognitive impairment (i.e., unable to understand the purpose of training and/or to provide informed consent), as determined by the healthcare team; and/or
- Acute illness (e.g., vomiting, fever), body mass >150 kg (exceeds safety harness system weight limits), colostomy bags, indwelling catheter, infection, pressure sore on pelvis or trunk.

Additionally, for safety reasons, participants who were unable to walk short distances (~10 m) without a gait aid were excluded from the assessment of balance reactions on a movable platform (described below). Eligibility was confirmed using information in participants’ hospital charts, by consulting members of participants’ healthcare team, and by consulting participants directly.

### Interventions

Participants were allocated to one of three groups: one (RBT1), three (RBT3), or six (RBT6), 45-minute RBT sessions. Previous research reported improved reactive balance control and reduced fall risk post-stroke after a single session of RBT, with 11-30 perturbations in the session.^9,10^ In another study, where volume of RBT varied among participants, reduced fall risk was observed after a median of 6 RBT sessions (180 minutes of training).^17^ Therefore, one, three, and six 45-minute sessions were chosen to cover the breadth of possible effective RBT volumes from previous studies. We initially planned for RBT to replace a portion of participants’ regular physiotherapy, so that the total amount of physical rehabilitation would not be affected by study participation, and would be approximately equal for the three groups. However, we were only able to enrol one participant in the first year of recruiting with this plan in place. Clinicians at our facility suggested that recruiting would be more successful if the intervention was offered outside of, rather than replacing, routine care. The protocol was subsequently amended to offer 6 sessions of balance and mobility training to all participants, with either one, three, or six of these comprising RBT (per group allocation), and the remainder including conventional balance and mobility exercises adapted from the Keep Moving with Stroke program.^18^ All six sessions occurred over 2-3 weeks.

A research physiotherapist oversaw the training sessions. Each 45-minute RBT session included approximately 60 perturbations. Rest breaks were scheduled into each session, and were provided when requested by participants. Training strategies were individualized to each participant, based on their balance impairments and rehabilitation goals.^4,19^ RBT sessions included multi-directional ‘internal’ and ‘external’ balance perturbations. Internal perturbations were achieved by asking the participant to complete tasks that challenge balance control, such that they lost balance when attempting to perform the task, and needed to execute a reactive step to recover balance. External perturbations were delivered manually using a push or pull from the physiotherapist while the participant was either standing still or doing a voluntary task, like marching on the spot; when the physiotherapist was positioned behind the participant, the direction and timing of the push or pull could be unpredictable to the participant. As participants improved their reactive balance control, difficulty was increased by shifting task requirements along a continuum from stable to mobile, and from predictable to unpredictable, and by increasing perturbation magnitude (i.e., by increasing the force of the push/pull) or imposing sensory or environmental challenges.^19^

### Outcomes

To assess feasibility of the trial, we documented rates of accrual (i.e., number of patients approached to participate in the study versus the number who provided consent), number of training sessions attended/missed, reasons for missed sessions, rate of missing data for the outcomes described below, and rate of withdrawal from the study.

Demographic, stroke information, and medical history were extracted from participants’ hospital charts or obtained directly from participants. Participants completed a questionnaire at baseline that asked about their social supports, employment, familial responsibilities, living situation etc., which are factors that could influence fall risk; items in this questionnaire were adapted from the Canadian Longitudinal Study on Aging.^20^(p201) The National Institutes of Health Stroke Scale (NIH-SS)^21^ and Chedoke-McMaster Stroke Assessment (CMSA;^22^ foot and leg) were scored at study enrolment.

Clinical assessments were scored by a blinded research assistant at three time points: 1) study enrolment; 2) post-training; and 3) 6 months post-training. Tests were: the; the mini-Balance Evaluation Systems Test (mini-BEST);^23^ the Activities-specific Balance Confidence (ABC) scale;^24^ and balance reactions following unpredictable and novel perturbations.

To assess balance reactions, participants completed 10 walking trials on a movable platform.

There were four force plates (model BP11971197-2000, Advanced Mechanical Technology, Inc., Watertown, Massachusetts, USA) embedded in the movable platform. On two trials, the platform suddenly moved forward on heel strike (i.e., when one of the force plates was loaded) to trigger a slip-like perturbation.^25,26^ On two other trials, the platform suddenly moved backward on toe-off (i.e., when one of the force plates was unloaded) to trigger a trip-like perturbation. Each slip or trip trial was triggered on heel-strike or toe-off, respectively, of each of the affected and unaffected limbs, either ‘early’ or ‘late’ in the walk based on which force plate was loaded/unloaded. The perturbation waveform consisted of a 300 ms square-wave acceleration, followed immediately by 300 ms deceleration.^25,26^ The peak platform acceleration was 1.0 m/s^2^ for participants who ordinarily use a gait aid for ambulation, and 1.5 m/s^2^ for all other participants. The platform only moved during these four trials; the remaining 6 trials consisted unperturbed walking. The perturbed and unperturbed walking trials were presented in a pseudo-random order to ensure that participants could not predict the timing (‘early’ vs ‘late’), direction (slip or trip), or perturbed limb (affected or unaffected) for these trials. The perturbation order was counterbalanced across participants, and each participant completed perturbations in the same order as their baseline visit on subsequent visits. Prior to completing the block of 10 perturbed and unperturbed walking trials, participants completed 3 unperturbed walking trials to familiarize them with walking on the platform; participants were told that the platform would not move during these familiarization trials. Participants were outfitted with reflective markers bilaterally on the tip of big toe, 2nd metatarsal-phalangeal joint, 3rd metatarsal phalangeal joint, calcaneus, lateral malleolus, medial malleolus, lateral femoral condyle, medial femoral condyle, greater trochanter, acromion process, lateral humeral epicondyle, medial humeral epicondyle, radial styloid process, and ulnar styloid process. Marker clusters were also placed on the moving platform and on the participants’ head, upper arms, thighs, shanks, chest, and pelvis. The locations of the reflective markers in space were recorded using a 9-12 camera motion capture system (Vicon Mx 40+ and MX F20 or Vicon Vero 2.2, Vicon Motion Capture Systems Ltd., Oxford, UK). An accelerometer (Series 7523A, Dynamic Transducer and Systems, Chatsworth, California, USA) placed on the platform recorded platform acceleration.

Participants were asked to report falls (“an event that results in a person coming to rest unintentionally on the ground or other lower level”^27^ in the 6 months post-training. Participants were provided with stamped, addressed postcards to mail to the research team every 2 weeks for 6 months post-training. Postcards contained a calendar, on which participants recorded falls. The blinded research assistant called participants who did not return the postcard to determine if any falls occurred. The research assistant contacted participants reporting a fall to complete a short questionnaire determining the cause and consequences of the fall. This method is considered the ‘gold standard’ for fall reporting.^28^ Participants also reported physical activities using the Physical Activity Scale for Individuals with Physical Disabilities (PASIPD),^29^ and participation in daily life using the Subjective Index of Physical and Social Outcome (SIPSO)^30^ at 2-, 4- and 6-months post-training.

### Harms

The physiotherapist observed and monitored participants for adverse events during the sessions (e.g., injuries, pain, expressions of fear/anxiety), and asked participants to report any adverse events at each session. Adverse events were documented using an adverse event reporting form (prior to April 2022) in our institution’s health information system, using the Common Terminology Criteria for Adverse Events (version 5.0; CTCAE; after April 2022).^31^ Any adverse events not included in the CTCAE were documented separately using the adverse event reporting form. Documentation included a narrative description of the adverse event, adverse event category, severity, whether the adverse event was attributed to the study intervention/procedures, and impact of the adverse event on the study. The physiotherapist ensured that participants received appropriate care if immediate attention was required (e.g., injuries during training sessions). The physiotherapist could modify the intervention in response to minor adverse events (e.g., reducing the number of perturbations due to fatigue, mild pain, or fear/anxiety). All adverse events were reviewed by the principal investigator.

### Sample size

We aimed to recruit 12 participants per group (36 participants total), as recommended for pilot studies.^32^

### Randomization

Participants were assigned using blocked randomization to one of the three different volumes of RBT (block size: 6). The random allocation sequence was computer generated. Blocked randomization was used to ensure equal numbers allocated to each group. Group allocation was performed centrally by the principal investigator, who was not involved in recruiting, scoring assessments, or administering the interventions (i.e., concealed allocation). Personnel who enrolled participants did not have access to the random allocation sequence.

### Blinding

Outcome measures were obtained by a research assistant who was blinded to group allocation. Participants cannot be blinded to group allocation. Data analysis was conducted by an individual who was not blinded to group allocation.

### Data processing

For the slip and trip trials, kinematic data were filtered at 6 Hz, and force plate and accelerometer data were filtered at 10 Hz using a 2^nd^ order zero phase lag Butterworth filter. Whole-body centre of mass (COM) was calculated using an 11-segment model (head and trunk, forearms and hands, upper arms, thighs, shanks, and feet).^33^ Perturbation onset time was determined from the accelerometer and acceleration of floor markers, and was the time when acceleration exceeded 0.1m/s^2^. Recovery foot off and contact times were the times when vertical force recorded by the force plate under the recovery limb was <1% body weight and >2% body weight, respectively. Foot off and contact times were visually inspected and corrected, if necessary (e.g., in the event of noise in the signal). Biomechanical stability at recovery foot contact was measured using an established method.^34^ Moving platform velocity was subtracted from COM velocity, and COM velocity was then normalized by dividing by the square root of gravitational acceleration multiplied by participant height. COM position was normalized to foot length. Stability was calculated as the perpendicular distance between the COM position-velocity curve and pre-defined anterior (trip) and posterior (slip) stability boundaries^35^ of the recovery limb. For trips, the sign of the stability metric was reversed so that, for both slips and trips, a negative value means that the COM position-velocity is behind (slips) or in front of (trips) the recovery. Thus, higher stability values indicate a more stable response.

### Statistical methods

The primary outcome in the larger trial will be rate of falls in daily life. To determine the optimal sample size for the larger trial, we used the rate of falls (number of falls per person-year) in the one-session group, reported over the 6-months post-discharge, and a clinically meaningful 30% reduction in fall rates.^36^ We then used the accrual rate (number of participants recruited per month) and proportion of participants who withdrew from the study to estimate how long it will take to achieve the target sample size in the larger trial. To assess feasibility of outcome collection, we calculated completeness of data collection for outcomes at each time point (i.e., slip- and trip-like perturbations, mini-BEST, ABC scale, PASIPD, and SIPSO). Between-group effect sizes were also calculated for all outcome measures. To assess feasibility of training with the prescribed volume, we calculated the mean number of sessions attended, and the mean number of perturbations per session.

The larger trial will compare one session of RBT with a higher volume. We used the reactive control sub-scale of the mini-BEST as a measure of effect of RBT on reactive balance control in each group. We calculated the pre-to-post training effect sizes for this sub-scale for each group. The minimum detectable change for the total mini-BEST score in people with stroke is 3 points^37^ (i.e., ~10% of the maximum score). The minimum detectable change for individual sub-scales have not been established, but we assumed that this is 10% of the maximum score for the subscale (i.e., 0.6 points), and compared between-group differences in pre-to-post-training effect sizes to this minimum detectable change.

## RESULTS

### Recruitment

Participants were enrolled from 20 August 2020 to 18 March 2025; the final assessment was completed on 22 October 2025. The trial ended when the target sample size had been recruited. Thirty-six participants were eligible and agreed to participate in the study. One participant (RBT1) had a change in health status that made them no longer eligible for the study after randomization, but prior to initiating training; therefore, analysis of data related to training is based on 35 participants (Research Question 4). One participant (RBT6) completed the intervention but withdrew prior to completing any assessments; therefore, analysis of secondary outcome data is based on 34 participants (Research Questions 3 and 5). One participant (RBT6) withdrew after completing the post-training data collection without completing any falls monitoring, and four participants (three RBT1 and one RBT6) declined any falls reporting; therefore, analysis of falls data is based on 30 participants (Research Questions 1 and 2; Figure 1). Participant characteristics are shown in Table 1.

**Table 1:** Participant characteristics. Values are means with standard deviations in parentheses for continuous or ordinal variables, and counts for categorical variables.

|  | **RBT1**<br>**(n=12)** | **RBT3**<br>**(n=12)** | **RBT6**<br>**(n=12)** |
| --- | --- | --- | --- |
| Age (years) | 63.8 (15.1) | 59.8 (15.0) | 56.4 (20.9) |
| Sex (number) |  |  |  |
| Female | 4 | 5 | 3 |
| Male | 8 | 7 | 9 |
| Height (m) | 1.67 (0.09) | 1.71 (0.10) | 1.70 (0.09) |
| Mass (kg) | 81.0 (19.4) | 78.9 (17.5) | 78.4 (17.8) |
| Time post-stroke (days) | 136 (45) | 104 (50) | 119 (43) |
| Stroke type (number) |  |  |  |
| Ischemic | 5 | 11 | 8 |
| Lacune | 5 | 1 | 3 |
| Hemorrhagic | 1 | 0 | 0 |
| Transient ischemic attack | 1 | 0 | 1 |
| Affected hemisphere (number) |  |  |  |
| Left | 6 | 1 | 4 |
| Right | 5 | 10 | 7 |
| Both | 1 | 1 | 1 |
| More affected side (number) |  |  |  |
| Left | 5 | 11 | 9 |
| Right | 7 | 1 | 3 |
| CMSA leg (score) | 5.2 (0.9) | 5.7 (0.7) | 5.8 (0.6) |
| CMSA foot (score) | 5.1 (1.4) | 5.3 (1.2) | 6.0 (1.0) |
| NIH-SS (score) | 0.8 (1.2) | 0.5 (0.9) | 0.8 (1.2) |

CMSA=Chedoke-McMaster Stroke Assessment, NIH-SS=National Institutes of Health Stroke Scale

**Figure 1:**
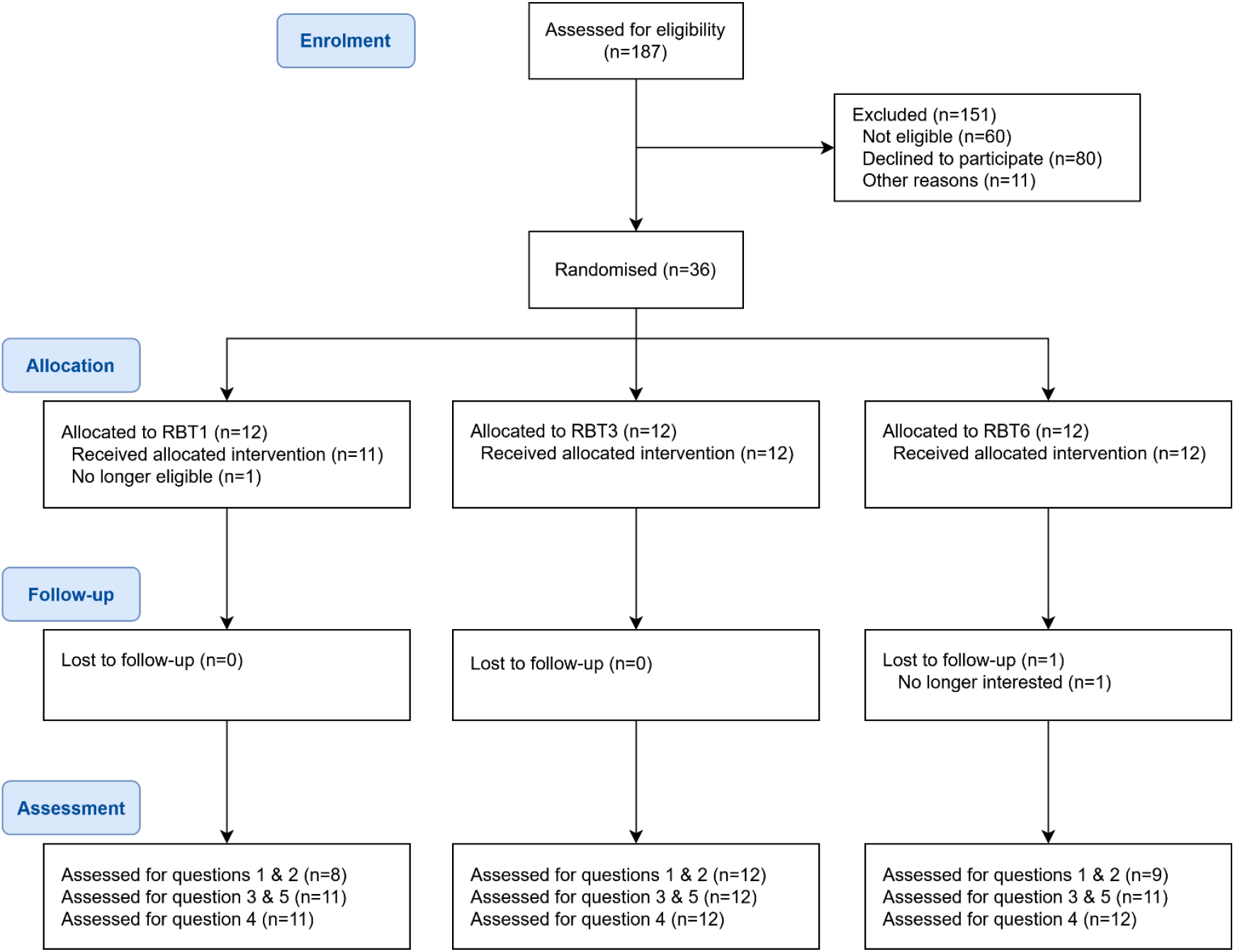
Participant flow.

### Research Questions 1 & 2: sample size and feasibility of recruiting

Fifteen falls were reported by nine participants during the 6-month follow-up period. Falls data are presented in Table 2. Using a mean of 1.25 falls per person-year for the control group, estimated dispersion of 0.26, and a 30% reduction in falls for the higher-volume group, the total sample size for larger trial would be 625. Increasing the monitoring duration to 1 year (assuming an actual mean monitoring duration of 11 months^4^), the required total sample size is 326. Accounting for 20% missing data, we would need to recruit 782 participants with a 6-month follow-up or 408 with a 1-year follow-up for a larger trial. From early 2022 onwards, when COVID-19 pandemic restrictions had been fully lifted, we recruited ~11 participants per year into the pilot study. Extrapolating this rate of recruiting, it would not be feasible to recruit from a single site for a larger trial using falls as the primary outcome.

**Table 2:**
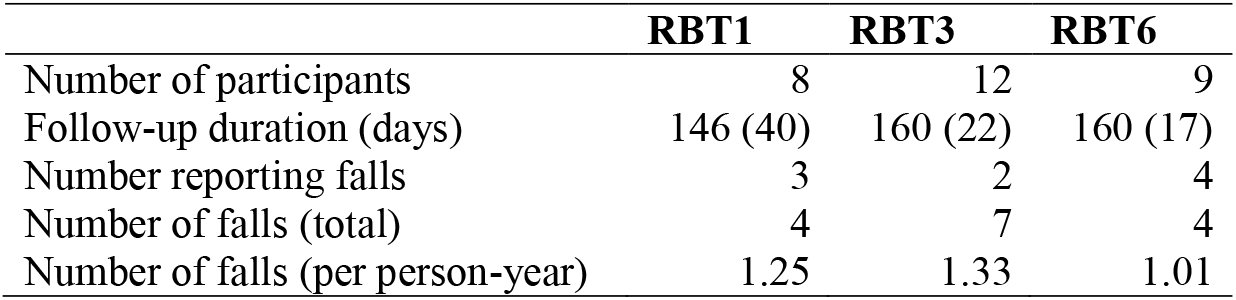
Falls data. Follow-up duration is presented as mean with standard deviation in parentheses; all other variables are total counts.

|  | **RBT1** | **RBT3** | **RBT6** |
| --- | --- | --- | --- |
| Number of participants | 8 | 12 | 9 |
| Follow-up duration (days) | 146 (40) | 160 (22) | 160 (17) |
| Number reporting falls | 3 | 2 | 4 |
| Number of falls (total) | 4 | 7 | 4 |
| Number of falls (per person-year) | 1.25 | 1.33 | 1.01 |

### Research Question 3: feasibility of outcome assessment

Two participants (RBT1) withdrew from the study prior to the 2-month follow-up time point, one participant (RBT1) withdrew between the 2- and 4-month time points, and two additional participants (RBT1) withdrew prior to the 6-month follow-up time point. One participant (RBT1) declined to complete the post-training assessment, six participants (two in each group) declined to complete the 2-month follow-up assessment, and five participants (three RBT1 and one in each of RBT3 and RBT6) declined to complete the 4-month follow-up assessment. Assessments were typically declined due to participant illness or vacation. Two participants (RBT1 at baseline and RBT6 post-training) declined to complete the ABC scale, but completed all other assessments at those time points. Consequently, data were available for >80% of participants for all functional and questionnaire outcomes, except the PASIPD and SIPSO at the 2- and 4-month follow-up time points (Table 3).

**Table 3:** Missing data.

|  | **RBT1**<br>**(n=11)** | **RBT3**<br>**(n=12)** | **RBT6**<br>**(n=11)** | **Total**<br>**(n=34)** | **% complete** |
| --- | --- | --- | --- | --- | --- |
| **Falls** |  |  |  |  |  |
| Complete | 8 | 12 | 9 | 29 | 85 |
| Withdrawn | 0 | 0 | 1 | 1 |  |
| Declined | 3 | 0 | 1 | 4 |  |
| **Baseline** |  |  |  |  |  |
| Mini-BEST |  |  |  |  |  |
| Complete | 11 | 12 | 11 | 34 | 100 |
| ABC |  |  |  |  |  |
| Complete | 10 | 12 | 11 | 33 | 97 |
| Declined | 1 | 0 | 0 | 1 |  |
| **Post-training** |  |  |  |  |  |
| Mini-BEST |  |  |  |  |  |
| Complete | 10 | 12 | 11 | 33 | 97 |
| Time-point missed | 1 | 0 | 0 | 1 |  |
| ABC |  |  |  |  |  |
| Complete | 10 | 12 | 10 | 32 | 94 |
| Time-point missed | 1 | 0 | 0 | 1 |  |
| Declined | 0 | 0 | 1 | 1 |  |
| **2 months post-training** |  |  |  |  |  |
| PASIPD & SIPSO |  |  |  |  |  |
| Complete | 7 | 10 | 8 | 25 | 74 |
| Withdrawn | 2 | 0 | 1 | 3 |  |
| Time-point missed | 2 | 2 | 2 | 6 |  |
| **4 months post-training** |  |  |  |  |  |
| PASIPD & SIPSO |  |  |  |  |  |
| Complete | 5 | 11 | 9 | 25 | 74 |
| Withdrawn | 3 | 0 | 1 | 4 |  |
| Time-point missed | 3 | 1 | 1 | 5 |  |
| **6 months post-training** |  |  |  |  |  |
| Mini-BEST, ABC, PASIPD & SIPSO |  |  |  |  |  |
| Complete | 6 | 12 | 10 | 28 | 82 |
| Withdrawn | 5 | 0 | 1 | 6 |  |

ABC=Activities Specific Balance Confidence scale; mini-BEST=mini-Balance Evaluation Systems Test; PASIPD=Physical Activity Scale for Individuals with Physical Disabilities; SIPSO=Subjective Index of Physical and Social Outcome

Two participants (RBT3) declined all moving platform perturbations at baseline, and two participants (RBT6) completed only one moving platform perturbation at baseline. Two of these participants (RBT3) were able to complete all moving platform perturbations post-training and one (RBT6) completed one additional perturbation post-training. All participants who completed the 6-month follow-up assessment completed all perturbations. Therefore, each type of perturbation trial was completed for >80% of participants at each time point. However, 21 perturbation trials total could not be included in analysis due to the perturbation being triggered at the wrong time in the gait cycle; for 13 trials, participants did not ‘cleanly’ load or unload the force plate (e.g., contacted the triggering force plate on toe contact instead of heel contact), for 4 trials the wrong trial was selected due to human error, and 4 trials (all unaffected trip trials) were affected by a programming error (this error was corrected early in the study and only affected the first few participants). Therefore, <80% of trials were available for analysis for affected slips at the 6-month follow-up (27/34), unaffected slips at baseline (27/34) and 6-month follow-up (25/34), and unaffected trips at 6-month follow-up (27/34).

### Research Question 4: intervention feasibility

All RBT1 participants completed one session of RBT, 11/12 RBT3 participants completed three RBT sessions (one participant completed only 2 sessions), and 10/12 RBT6 participants completed six RBT sessions (2 participants completed only 4 sessions). Therefore, the mean number of RBT sessions/group was 1 (RBT1), 2.9 (RBT3), and 5.7 (RBT6). As noted above, the first participant (RBT3) was enrolled prior to amending the protocol to include a control intervention for the remainder of the training sessions, and was therefore only offered 3 balance training sessions, total. Four other participants (one RBT1, one RBT3, and two RBT6) attended 4-5 total balance training sessions.

Therefore, 31/35 participants (88.6%) completed the number of RBT and additional balance training sessions dictated by the protocol, and the mean number of sessions attended was 96.7% of prescribed.

The average number of perturbations per session was 64.5 (standard deviation: 15.1) for RBT1, 59.9 (standard deviation: 13.3) for RBT3, and 51.9 (standard deviation: 13.8) for RBT6. For two participants (both RBT6), the physiotherapist noted that participants’ reactive balance control was very good, and that it was difficult to challenge them sufficiently to evoke multi-step reactions. For two participants (one RBT1 and one RBT6), the physiotherapist felt that the volume of RBT was too low, and that those participants would likely benefit from more sessions.

### Research Question 5: effect sizes for secondary outcomes

Group means and effect sizes are presented in Tables 4 and 5. The mean improvement in mini-BEST reactive score from pre-to post-training was 0.4, 1.0 and 0.7 out of possible 6 points for RBT1, RBT3, and RBT6, respectively.

**Table 4:** Physical activity and quality of life post-training. Values presented are group means with standard deviations in parentheses at each time point. Group differences are presented as means with 95% confidence intervals in brackets.

|  | RBT1 |  | RBT3 |  | RBT6 |  | Group differences |  |
| --- | --- | --- | --- | --- | --- | --- | --- | --- |
|  | N | Estimate | N | Estimate | N | Estimate | RBT3 – RBT1 | RBT6 – RBT1 |
| **2 months** |  |  |  |  |  |  |  |  |
| PASIPD (score) | 7 | 19.7 (18.6) | 10 | 15.5 (9.5) | 8 | 13.8 (8.9) | -4.2 [-10.8, 2.4] | -5.9 [-13.1, 1.3] |
| SIPSO (score) | 7 | 30.9 (4.1) | 10 | 30.5 (5.4) | 8 | 30.9 (5.5) | -0.1 [-2.4, 2.2] | 0 [-2.5, 2.5] |
| **4 months** |  |  |  |  |  |  |  |  |
| PASIPD (score) | 5 | 14.2 (5.2) | 11 | 17.4 (9.0) | 9 | 10.9 (6.7) | 3.2 [-0.8, 7.2] | -3.3 [-6.6, 0.0] |
| SIPSO (score) | 5 | 30.6 (3.8) | 11 | 31.7 (5.0) | 9 | 30.8 (5.3) | 1.6 [-0.7, 3.9] | 0.2 [-2.3, 2.7] |
| **6 months** |  |  |  |  |  |  |  |  |
| PASIPD (score) | 6 | 16.7 (3.6) | 12 | 17.0 (7.5) | 10 | 13.5 (10.2) | 0.3 [-2.7, 3.3] | -3.3 [-7.4, 0.9] |
| SIPSO (score) | 6 | 32.5 (4.0) | 12 | 33.6 (3.2) | 10 | 28.2 (8.1) | 1.0 [-0.6, 2.6] | -4.3 [-7.7, -0.9] |

**Table 5:** Functional balance, balance confidence, and stability pre- and post-training. Values presented are group means with standard deviations in parentheses at each time point. Time and group differences are presented as means with 95% confidence intervals in brackets.

|  | RBT1 |  |  |  | RBT3 |  |  |  | RBT6 |  |  |  | Group differences<sup>†</sup> |  |
| --- | --- | --- | --- | --- | --- | --- | --- | --- | --- | --- | --- | --- | --- | --- |
|  | N | Estimate | N | Δ time<sup>*</sup> | N | Estimate | N | Δ time<sup>*</sup> | N | Estimate | N | Δ time<sup>*</sup> | RBT3-RBT1 | RBT6-RBT1 |
| **Mini-BEST total score (maximum score=28)** |  |  |  |  |  |  |  |  |  |  |  |  |  |  |
| Pre | 12 | 22.3 (2.6) |  |  | 12 | 22.4 (2.3) |  |  | 12 | 23.0 (2.8) |  |  |  |  |
| Post | 10 | 23.1 (2.4) | 10 | 0.5 [-0.8, 1.8] | 12 | 23.9 (3.1) | 12 | 1.5 [0.4, 2.6] | 11 | 23.1 (3.4) | 11 | 0.2 [-1.2, 1.6] | 1.0 [0.1, 1.9] | -0.3 [-1.3, 0.7] |
| 6 months | 6 | 24.0 (1.7) | 6 | 0.2 [-2.6, 2.9] | 12 | 23.6 (2.7) | 12 | 1.2 [0.2, 2.2] | 10 | 23.2 (3.0) | 10 | -0.3 [-1.4, 0.8] | 1.0 [-0.1, 2.1] | -0.5 [-1.7, 0.8] |
| **Mini-BEST reactive score (maximum score=6)** |  |  |  |  |  |  |  |  |  |  |  |  |  |  |
| Pre | 12 | 3.8 (1.4) |  |  | 12 | 3.4 (1.3) |  |  | 12 | 3.8 (1.3) |  |  |  |  |
| Post | 10 | 4.4 (1.2) | 10 | 0.4 [-0.6, 1.4] | 12 | 4.4 (1.0) | 12 | 1.0 [0.4, 1.6] | 11 | 4.5 (1.1) | 11 | 0.7 [0.1, 1.3] | 0.6 [0.04, 1.2] | 0.3 [-0.3, 0.9] |
| 6 months | 6 | 4.5 (1.4) | 6 | 0.0 [-1.7, 1.7] | 12 | 4.2 (1.0) | 12 | 0.8 [-0.2, 1.7] | 10 | 4.1 (0.9) | 10 | 0.1 [-0.4, 0.6] | 0.8 [-0.1, 1.6] | 0.1 [-0.6, 0.8] |
| **ABC score (%)** |  |  |  |  |  |  |  |  |  |  |  |  |  |  |
| Pre | 11 | 78.7 (13.2) |  |  | 12 | 73.3 (22.2) |  |  | 12 | 65.6 (21.1) |  |  |  |  |
| Post | 10 | 85.4 (12.4) | 9 | 7.5 [3.0, 12.0] | 12 | 83.1 (13.1) | 12 | 9.8 [3.8, 15.8] | 10 | 74.9 (16.8) | 10 | 4.9 [-1.2, 11.1] | 2.3 [-1.6, 6.3] | -2.5 [-6.5, 1.4] |
| 6 months | 6 | 90.4 (10.5) | 6 | 4.2 [-2.4, 10.8] | 12 | 83.5 (14.9) | 12 | 10.3 [2.9, 17.6] | 10 | 72.7 (29.1) | 10 | 4.9 [-5.9, 15.7] | 6.1 [0.7, 11.5] | 0.7 [-6.5, 8.0] |
| **Affected slip stability<sup>‡</sup>** |  |  |  |  |  |  |  |  |  |  |  |  |  |  |
| Pre | 10 | 0.60 (0.14) |  |  | 10 | 0.65 (0.28) |  |  | 9 | 0.58 (0.25) |  |  |  |  |
| Post | 9 | 0.58 (0.16) | 9 | -0.02 [-0.07, 0.03] | 12 | 0.70 (0.18) | 10 | 0.08 [-0.03, 0.19] | 7 | 0.55 (0.26) | 7 | 0.01 [-0.06, 0.08] | 0.10 [0.04, 0.16] | 0.03 [-0.01, 0.07] |
| 6 months | 5 | 0.69 (0.15) | 4 | -0.01 [-0.07, 0.05] | 12 | 0.72 (0.18) | 10 | 0.11 [-0.06, 0.27] | 10 | 0.60 (0.15) | 9 | 0.06 [-0.09, 0.21] | 0.12 [0.001, 0.24] | 0.07 [-0.01, 0.16] |
| **Unaffected slip stability<sup>‡</sup>** |  |  |  |  |  |  |  |  |  |  |  |  |  |  |
| Pre | 11 | 0.63 (0.16) |  |  | 6 | 0.73 (0.14) |  |  | 11 | 0.49 (0.25) |  |  |  |  |
| Post | 9 | 0.68 (0.16) | 9 | 0.02 [-0.04, 0.08] | 11 | 0.78 (0.16) | 6 | 0.07 [-0.03, 0.18] | 10 | 0.59 (0.25) | 10 | 0.07 [-0.01, 0.15] | 0.05 [-0.01, 0.11] | 0.05 [-0.01, 0.11] |
| 6 months | 6 | 0.68 (0.19) | 6 | 0.00 [-0.15, 0.15] | 9 | 0.78 (0.18) | 5 | 0.05 [-0.14, 0.24] | 10 | 0.64 (0.18) | 10 | 0.12 [0.02, 0.22] | 0.05 [-0.07, 0.17] | 0.12 [0.04, 0.21] |
| **Affected trip stability<sup>‡</sup>** |  |  |  |  |  |  |  |  |  |  |  |  |  |  |
| Pre | 11 | 0.44 (0.14) |  |  | 9 | 0.49 (0.13) |  |  | 11 | 0.40 (0.07) |  |  |  |  |
| Post | 10 | 0.43 (0.12) | 10 | 0.00 [-0.04, 0.04] | 11 | 0.51 (0.13) | 8 | 0.05 [-0.03, 0.14] | 10 | 0.43 (0.10) | 10 | 0.01 [-0.06, 0.08] | 0.06 [0.01, 0.10] | 0.02 [-0.03, 0.06] |
| 6 months | 6 | 0.47 (0.16) | 6 | -0.03 [-0.08, 0.03] | 12 | 0.56 (0.12) | 9 | 0.09 [0.01, 0.18] | 10 | 0.44 (0.16) | 9 | 0.08 [0.04, 0.11] | 0.12 [0.06, 0.17] | 0.10 [0.07, 0.13] |
| **Unaffected trip stability<sup>‡</sup>** |  |  |  |  |  |  |  |  |  |  |  |  |  |  |
| Pre | 12 | 0.40 (0.10) |  |  | 10 | 0.39 (0.12) |  |  | 9 | 0.42 (0.11) |  |  |  |  |
| Post | 10 | 0.42 (0.18) | 10 | 0.01 [-0.10, 0.12] | 11 | 0.42 (0.09) | 9 | 0.05 [-0.03, 0.12] | 10 | 0.40 (0.17) | 8 | 0.04 [-0.001, 0.08] | 0.03 [-0.03, 0.10] | 0.03 [-0.04, 0.09] |
| 6 months | 7 | 0.32 (0.34) | 7 | -0.10 [-0.39, 0.18] | 12 | 0.47 (0.12) | 10 | 0.10 [0.01, 0.19] | 9 | 0.48 (0.06) | 8 | 0.07 [0.04, 0.11] | 0.20 [0.07, 0.33] | 0.18 [0.06, 0.30] |

\*Δ time is the difference from baseline, only including participants who had data available for both time points; <sup>†</sup>The difference in Δ time between each group and RBT1; <sup>‡</sup>Unitless measure

### Harms

Nine adverse events that were deemed possibly, probably, or definitely related to the intervention were experienced by six participants. These adverse events were joint pain (4 events from 1 participant in RBT6, and 1 event from 1 participant in RBT6), headache (1 event from 1 participant in RBT3), back pain (1 event from 1 participant in RBT6), non-cardiac chest pain (1 event from 1 participant in RBT3), and muscle cramp (1 event from 1 participant in RBT1).

### Ancillary analyses

Because the estimated sample size for the larger trial with falls at the primary outcome was so large, we also estimated the sample size with the mini-BEST reactive score and stability measures. We used 0.6 as a clinically meaningful training effect for the mini-BEST reactive score. Clinically meaningful effect sizes for stability measures are not available. However, one study reported change in stability of approximately 0.06 after conventional balance training (i.e., not focused on improving reactive balance control) among people with stroke.^38^ Assuming a minimum of twice as much change with RBT, we used an effect size of 0.12 for stability measures. Using sample size calculation for analysis of covariance^39^ (i.e., controlling for the pre-training value), observed standard deviations and rates of missing data from this pilot study, we calculated total sample sizes of 30 to 166 (Table 6), with alpha of 0.05 and 80% power.

**Table 6:** Estimated sample sizes using alternative primary outcomes. The effect size is the mean difference between groups, and variance was estimated from the current study.

| Outcome | Effect size | Variance | Sample size | Missing data (%) | Total sample size |
| --- | --- | --- | --- | --- | --- |
| Mini-BEST reactive score | 0.6 | 1.69 | 152 | 8 | 166 |
| Affected slip stability | 0.12 | 0.0196 | 46 | 31 | 68 |
| Unaffected slip stability | 0.12 | 0.0121 | 30 | 33 | 46 |
| Affected trip stability | 0.12 | 0.0081 | 22 | 22 | 30 |
| Unaffected trip stability | 0.12 | 0.0225 | 54 | 25 | 72 |

## DISCUSSION

This study aimed to assess the feasibility of a randomized controlled trial to determine the optimal volume of RBT for people with sub-acute stroke. We determined that, using falls as a primary outcome, the larger trial would not be feasible at a single site. Using biomechanical stability following balance perturbations as a primary outcome resulted in more feasible sample size estimates.

We aimed to assess feasibility of the study intervention. We found that it was not feasible to implement a specific volume of RBT into routine out-patient rehabilitation, as patients were generally not willing to participate in a study where a portion of their therapy was replaced with the study intervention. When the protocol was amended to complete the intervention outside of routine care intervention, fidelity was high, with most participants completing the volume of RBT dictated by their group allocation. This finding agrees with other studies, where adherence to RBT interventions is generally high among people with stroke.^4,5,40,41^ Therefore, future studies conducted with people still enrolled in stroke rehabilitation should supplement, rather than replace, participants’ usual care. We also reported few adverse events, with the frequency and type of events being similar to those reported in other exercise studies. This agrees with previous studies demonstrating the safety of RBT.^3^

We also aimed to determine the intervention groups that should be included in a future, larger trial. Within-group effect sizes from pre-to post-training suggest that both RBT3 and RBT6 improved mini-BEST reactive scores, with the mean change being within 0.6 points of each other (i.e., a clinically meaningful effect size). Therefore, based on our *a priori* criteria,^42^ a future trial should include RBT1 and RBT3. Between-group effect sizes also suggest that RBT3 had a greater improvement in mini-BEST total and reactive scores, and affected slip and trip stability, than RBT1 from preto post-training. The possible benefit of six 45-minute sessions of RBT (135 minutes total) compared to one session is supported by a previous non-randomized study, where people with subacute stroke who completed a median of six 30-minute RBT sessions (180 minutes total) experienced fewer falls in daily life than those who did not complete RBT.^17^ There was no apparent benefit for RBT6 compared to RBT3, based on between-group effect sizes. This contrasts with another cross-sectional study showing a positive correlation between the volume of RBT completed and change in mini-BEST scores among people with chronic stroke.^11^ This difference in findings may be partially explained by the differing study populations; the current study included people with sub-acute stroke, which is a stage of stroke recovery marked by high potential for neuroplasticity.^43^ A lower volume of RBT may be sufficient to improve reactive balance control in this neuroplastic sub-acute stage compared to the chronic stage. Indeed, others have found that when the same volume of exercise is delivered in the chronic versus sub-acute stage of stroke recovery, greater gains are observed during the sub-acute stage.^44^ A meta-regression also found no evidence for greater efficacy of RBT (in terms of reactive balance control or fall risk) when more perturbations were completed during training.^6^ It is possible that a plateau is reached after a certain volume of RBT, above which any further benefits are not observed; this possibility is suggested by our findings. However, the optimal volume of RBT for different populations will need to be confirmed in future studies.

Our stability data suggest that participants improved stability following the slip and trip perturbations, indicating transfer of improvements in reactive balance control to a type of perturbation that was not included in training. Importantly, these gains were most apparent at the 6-month follow-up assessment. While it is possible that these improvements in stability may be due to repetition of the slip and trip tests, rather than transfer of learning, previous work suggests that experiencing a single slip or trip perturbation does not lead to large and lasting improvements in responses to the perturbations.^45,46^ Furthermore, our data show greater improvements in stability for RBT3 and RBT6 than RBT1, which again suggests transfer of learning among those participants who completed a higher volume of training.

### Limitations

Estimating sample size for a future trial using stability outcomes following balance perturbations as a primary outcome resulted in more feasible sample size estimates, despite a high rate of missing data for these outcomes (>20% missing). Data were primarily missing for stability measures due to perturbations triggered incorrectly when participants made ‘unclean’ contact with the force plate. This issue could be solved in future research by using more sophisticated perturbation triggering rules. The minimum clinically important difference or minimum detectable change has also not been established for these outcomes; we estimated the smallest effect size of interest^47^ based on a previous study of changes in stability after ‘conventional’ balance training,^38^ where reactive balance control would not be expected to improve. Effect sizes for the stability outcome in RBT studies post-stroke have been reported to be approximately 2-6 times larger than our estimated smallest effect size of interest.^9,48^ Clinically meaningful effect sizes for this outcome will need to be established in future work.

### Conclusions

The modified intervention delivery and outcome assessment were generally feasible. However, a larger trial using falls as an outcome would not be feasible at a single site. While using stability outcomes as a proxy for fall risk resulted in more feasible sample size calculations, clinically meaningful effects will need to be established for these outcomes.

## Data Availability

Data are not available publicly due to local privacy legislation.

## Notes

**Funding:** This study was supported by the Heart and Stroke Foundation Canadian Partnership for Stroke Recovery. We also acknowledge the support of the Toronto Rehabilitation Institute; equipment and space have been funded with grants from the Canada Foundation for Innovation, Ontario Innovation Trust, and the Ministry of Research and Innovation. These funding sources had no role in the design or execution of the study, analysis, interpretation of the data, or decision to submit results.

**Conflicts of interest:** None declared.

### Competing Interest Statement

The authors have declared no competing interest.

### Clinical Trial

clinicaltrials.gov, NCT04219696

### Clinical Protocols

https://doi.org/10.1136/bmjopen-2020-038073

### Author Declarations

Ethics committee of the University Health Network gave ethical approval for this work (Study ID: 19-6001)

